# From online data collection to identification of disease mechanisms: The IL-1ß, IL-6 and TNF-α cytokine triad is associated with post-acute sequelae of COVID-19 in a digital research cohort

**DOI:** 10.1101/2021.11.16.21266391

**Authors:** Christoph Schultheiß, Edith Willscher, Lisa Paschold, Cornelia Gottschick, Bianca Klee, Svenja-Sibylla Henkes, Lidia Bosurgi, Jochen Dutzmann, Daniel Sedding, Thomas Frese, Matthias Girndt, Jessica I. Höll, Michael Gekle, Rafael Mikolajczyk, Mascha Binder

## Abstract

Post-acute sequelae of COVID-19 (PASC) emerge as a global problem with unknown molecular drivers. In a digital epidemiology approach, we rapidly recruited 8,077 individuals out of 129,733 households in Halle (Saale) to the cohort study for digital health research in Germany (DigiHero). These responded to a basic questionnaire followed by a PASC-focused survey and blood sampling in case of prior positive SARS-CoV-2 testing in their household. The presented analysis is based on the first 318 DigiHero participants, the majority thereof after mild infections. PASC were reported in 67.8% of cases, consisted predominantly in fatigue, dyspnea and concentration deficit, persisted in 60% over the follow-up period of on average eight months and their resolution was unaffected by post-infection vaccination. PASC was not associated with post-COVID-19 autoantibodies, but with elevated levels of IL-1ß, IL-6 and TNF-α. Blood profiling and single-cell data from validation cohorts with early infection suggested the induction of these cytokines in COVID-19 lung pro-inflammatory macrophages creating a self-sustaining feedback loop. Our data indicate a long-lasting cytokine triad -potentially underlying PASC symptoms - to be driven by macrophage primed during infection. We demonstrate how the combination of digital epidemiology with selective biobanking can rapidly generate hints towards disease mechanisms.

## Introduction

Severe acute respiratory syndrome coronavirus 2 (SARS-CoV-2) is a new virus causing coronavirus disease 2019 (COVID-19) which has led to a health crisis of global scale[1, 2]. COVID-19 is now recognized as a multi-organ disease with considerable mortality in risk groups[3-5]. With a growing population of recovering patients, it became clear that in 32-87% of patients (including those with mild acute disease) health impairments persist beyond the acute phase of infection[6-10]. The most common definition of such post-acute sequelae of COVID-19 (PASC) is persistence of symptoms beyond four weeks[6, 7]. The clinical spectrum of PASC includes fatigue and exercise intolerance, brain fog, shortness of breath, joint pain, fever, sleep and anxiety disorders as well as gastrointestinal symptoms and palpitations[6-8]. Symptoms may persist for months and their severity can range from mild to debilitating. The immense numbers of COVID-19 survivors with post-infection disability that prevent these individuals to return to normal active life added another layer in this health crisis beyond the threat of exhausting intensive care unit capacities. Given 250 million SARS-CoV-2 infections counted globally until November 2021 by the WHO, the impact of PASC will likely be profound.

The pathophysiology of PASC is still largely unexplored[6, 8] and therefore targeted treatment approaches are lacking. While some of the delayed symptoms may be a consequence of virus-induced tissue injury affecting multiple organs[11, 12], another potential trigger has been proposed to result from persistent SARS-CoV-2 reservoirs. This hypothesis has been fueled by the observation that some infected patients do not rapidly clear the virus[13-16], which is in line with the observation that post-acute viral persistance is a relatively common feature of RNA viruses (e.g. Ebola and HCV) and has also been discussed in the context of chronic symptoms or reactivated disease[17]. Yet, direct evidence pointing to a role of such potential reservoirs in PASC as well as on the effects of their eradication e.g. via post-infection vaccination is currently lacking[18]. Another potential biological correlate of PASC may be autoimmune tissue damage. Already in the early phases of the pandemic, it became obvious that the SARS-CoV-2 virus shifts adaptive immunity towards autoreactivity[19, 20]. There is now a large body of evidence that diverse autoantibody classes are produced in acute COVID-19 as well as post-COVID-19 multisystem inflammatory syndrome in children[20-27]. Moreover, many reports suggest that patients may experience de novo or worsening of preexisting autoimmune conditions such as autoimmune cytopenias, Guillain-Barré syndrome or systemic lupus erythematodes[28, 29]. It remains elusive, however, if autoantibodies represent an inflammatory epiphenomenon or pathophysiologically contribute to PASC[30].

Quickly closing the knowledge gap on PASC pathophysiology is one of the current global priorities. Here, we show how the combination of digital epidemiology with selective biobanking can rapidly generate hints towards disease mechanisms. Using this approach, we rapidly identified and recruited a large cohort allowing dedicated analyses of biomaterial in a subsample of previously infected participants with or without PASC. Our analyzes provide evidence for a long-lasting cytokine signature consisting of elevated levels of IL-1ß, IL-6 and TNF-α that potentially underlie many of the clinical symptoms of PASC and that may derive from the macrophage compartment.

## Methods

### COVID-19 module within the DigiHero cohort study (Population-based cohort study for digital health research in Germany)

Until October 2021, 8,077 individuals took part in the digital cohort study DigiHero in the city of Halle (Saale), Germany. The recruitment was conducted in two waves and included mailed invitation to all 129,733 households in Halle as well as promotion via media. Of these, 919 individuals reported prior positive SARS-CoV-2 testing in their households and were thus invited – along with their household members – to take part in the COVID-19 module of the study. Figure 1 provides a flow-chart of the COVID-19 module within the DigiHero study. Until 9^th^ of October 2021, 318 individuals older than 14 years had been recruited to this module. All participants were interviewed with a questionnaire on the clinical course of COVID-19 and its sequelae as well as on vaccination status. The study was approved by the institutional review board (approval numbers 2020-076) and conducted in accordance with the ethical principles stated by the Declaration of Helsinki. Informed written consent was obtained from all participants or legal representatives. The collected plasma samples were isolated by centrifugation of whole blood for 15 min at 2,000xg, followed by centrifugation at 12,000xg for 10 min and stored at - 80°C.

**Figure 1.**
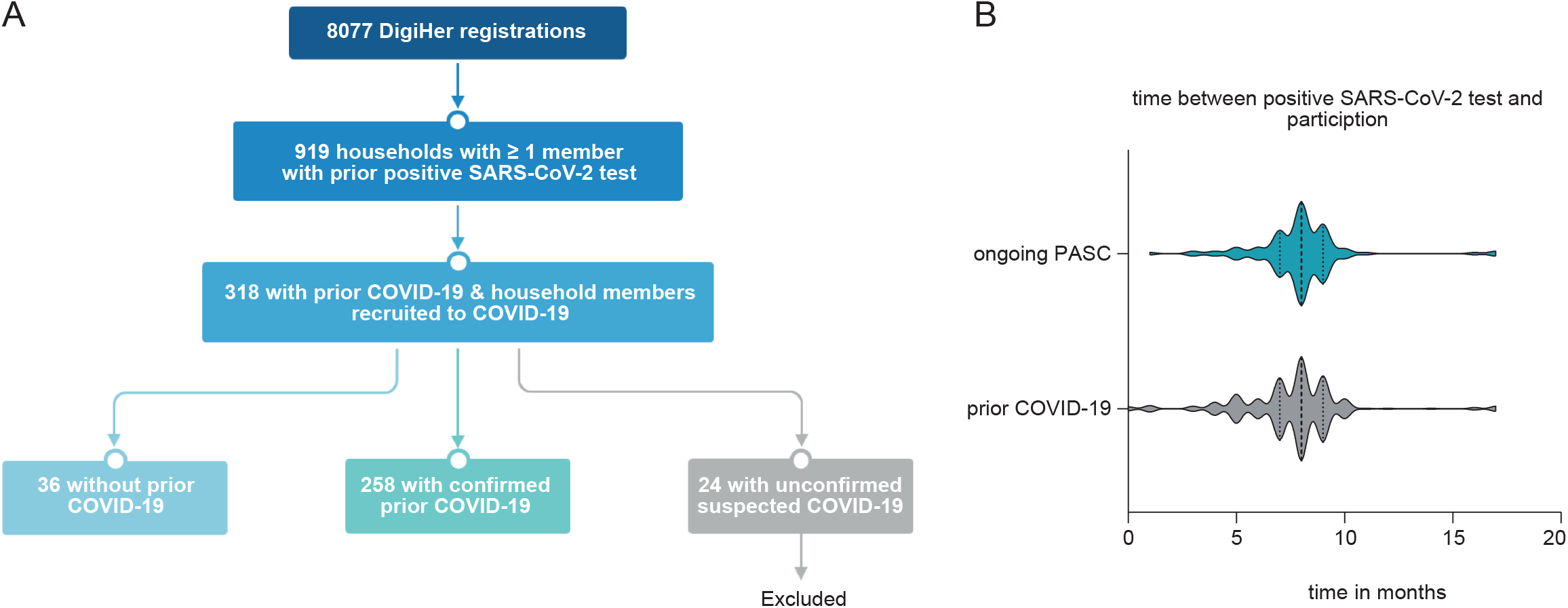
The COVID-19 module of the DigiHero study. (A) Flow-chart of the COVID-19 module of the DigiHero study. (B) Median time from positive PCR or antigen test to participation in the module for the no prior COVID-19 and ongoing PASC groups.

### Biological samples and data from the biobank of the Halle COVID cohort (HACO)

Plasma samples from acute bacterial pneumonia (n=5), acute COVID-19 (n=15 mild to moderate severity) and early post-acute COVID-19 (n=49 mild to moderate severity) were recovered from the biobank of the HACO study that recruited participants from April to December 2020. Informed written consent was obtained and the study was approved by the institutional review board (approval number 2020-039) and conducted in accordance with the ethical principles stated by the Declaration of Helsinki. The collected plasma samples were isolated as described above.

### SARS-CoV-2 antibody profiling

Antibodies against the S1 domain of the spike (S) protein and the nucleocapsid protein (NCP) of SARS-CoV-2 were determined by Anti-SARS-CoV-2-ELISA IgA/IgG and Anti-SARS-CoV-2-NCP-ELISA kits from Euroimmun (Lübeck, Germany). Readouts were performed at 450 nm using a Tecan Spectrophotometer SpectraFluor Plus (Tecan Group Ltd., Männedorf, Switzerland).

### Cytokine and autoantibody profiling

Cytokine plasma levels were measured using the LEGENDplex Human B Cell Panel (13 plex) and the Human Anti-Virus Response Panel (13-plex) (BioLegend) on a BD FACSCelesta flow cytometer. For autoantibody screens, the Rheumatoid Factor (detects IgG, IgA and IgM RFs), ANA (detects SS-A 60, SS-A 52, SS-B, RNP-70, Sm, RNP/Sm, Scl-70, centromere B and Jo-1 IgGs) and Anti-Phospholipid IgG/IgM (detects cardiolipin, phosphatidylserine, phosphatidylinositol, phosphoglycerides und β2-glycoprotein 1 IgGs/IgMs) kits from Orgentec (Mainz, Germany) were used.

### Single-cell transcriptome analyzes

Single-cell RNA sequencing data of three previously published datasets was analyzed in R (v 4.1.1) using the package Seurat (v 4.0.3). Dataset 1 contained bronchoalveolar lavage fluid (BAL-F) and blood cells from seven patients with SARS-CoV-2 infection and four patients with bacterial pneumonia[31]. Dataset 2 contained lung autopsy tissues from 16 deceased COVID-19 patients[32]. Dataset 3 contained peripheral blood mononuclear cells from seven hospitalized COVID-19 patients and six healthy controls[33]. Cytokine response scores of gene sets were calculated using the Seurat function *AddModuleScore*. The IL-1β set encompassed *IL1A, IL1B, CASP1, NLRP1, NLRP3, TLR7, FOSB, NFKBIZ, NFKB1*, the IL-6 set *IL6, CEBPD, NOTCH1, HES1, HES4, HEY1* and the TNF-α set *TNFSF10, TNFSF15, CLU, TNIP3, SIGLEC10, ENPP2, NKG7, TIMP1, CLEC5A* and *CCL7*[34].

### Statistical analysis

We used logistic regression to assess acute COVID-19 symptoms associated with PASC by fitting a generalized linear model with R function *glm*. Barplots, plasma level heatmap and all statistical analyses were performed using GraphPad Prism 8.3.1 (GraphPad Software, La Jolla, CA, USA). Differences in plasma cytokine levels were studied by unpaired t-test with Welch’s correction.

## Results

### Characteristics of participants in the DigiHero COVID-19 module

A total of 258 individuals with prior COVID-19 (as confirmed by a positive PCR or antigen test) and 36 presumably uninfected household members (no symptoms, no positive PCR or antigen test) participated in the module (Figure 1A). Twenty-four individuals with suspected infection due to symptoms were excluded from the analyzes presented in this manuscript due to lack of confirmed infection. Basic characteristics of the cohort are summarized in Table 1. More than 76.7% of the previously infected participants had COVID-19 or asymptomatic SARS-CoV-2 infections in Germany’s second wave. More than 80% of acute infections were rated mild to moderate by the participants. Median time from positive PCR or antigen test to participation in the module was 8 months (Figure 1B). More than 80% of participants had received at least one dose of a COVID-19 vaccine.

**Table 1:** Characteristics of participants in the DigiHero "COVID-19” module.

|  | all participants | participants with confirmed prior COVID-19 | Participants without prior COVID-19 |
| --- | --- | --- | --- |
| Nb of participants | 318 | 258 | 36 |
| Sex |  |  |  |
| Female | 192 (60.4%) | 161 (62.4%) | 22 (61.1%) |
| Male | 126 (39.6%) | 97 (37.6%) | 14 (38.9%) |
| Age (years) |  |  |  |
| Median age | 51.3 | 51.2 | 50 |
| Range | 15-83 | 15-83 | 17-81 |
| Timing of COVID-19 infections |  |  |  |
| Wave 1 (March 2020 -June 2020) |  | 7 (2.7%) |  |
| Wave 2 (July 2020 - Feb 2021) |  | 198 (76.7%) |  |
| Wave 3 (Feb 2021 - June 2021) |  | 45 (17.4%) |  |
| Wave 4 (from July 2021) |  | 8 (3.1%) |  |
| Self rated acute infection severity |  |  |  |
| asymptomatic |  | 15 (5.8%) |  |
| mild to moderate |  | 221 (85.7%) |  |
| severe |  | 22 (8.5%) |  |
| Hospitalization |  | 8 (3.1%) |  |
| Intensive care unit |  | 4 (1.5%) |  |
| Duration of symptoms |  |  |  |
| not evaluated |  | 12 (4.7%) |  |
| 0-4 weeks |  | 71 (27.5%) |  |
| 4-12 weeks |  | 30 (11.6%) |  |
| >12 weeks |  | 145 (56.2%) |  |
| Nb of SARS-COV-2 vaccinations |  |  |  |
| 0 |  | 62 (24%) | 6 (16.7%) |
| 1 |  | 137 (53.1%) | 0 |
| 2 |  | 59 (22.9%) | 30 (83.3%) |
| Type of SARS-CoV-2 vaccine |  | 196 |  |
| not evaluated |  | 25 (12.8%) | 1 (3.3%) |
| mRNA vaccine* |  | 128 (65.3%) |  |
| mRNA vaccine + mRNA vaccine |  | 29 (14.8%) | 24 (80%) |
| vector vaccine* |  | 7 (3.6%) |  |
| vector vaccine + vector vaccine |  | 3 (1.5%) | 1 (3.3%) |
| vector vaccine + mRNA vaccine |  | 4 (2%) | 4 (13.3%) |

\* mRNA vaccines: Comirnaty (BioNTech / Pfizer) or Spikevax (Moderna Biotech), vector vaccines: Vaxzevria (AstraZeneca) or Ad26.COV2-S (Johnson&Johnson)

### SARS-CoV-2 antibody results confirm infection/vaccination status information provided by participants

To assess validity of the provided information on infection and vaccination status, we performed ELISA testing for SARS-CoV-2 NCP and S1 antibodies (Figure 2A). These results confirmed prior evidence that patients vaccinated after natural infection achieved the highest S1 antibody levels followed by vaccinated, but uninfected individuals[35]. The lowest S1 antibody levels were detected in individuals after natural infection without subsequent vaccination and the few individuals that were neither previously infected, nor vaccinated. These data were well compatible with the information on infection/vaccination status provided by the participants except for two individuals that indicated no prior COVID-19 or vaccination despite elevated antibody levels. These two cases were excluded from further analyses.

**Figure 2.**
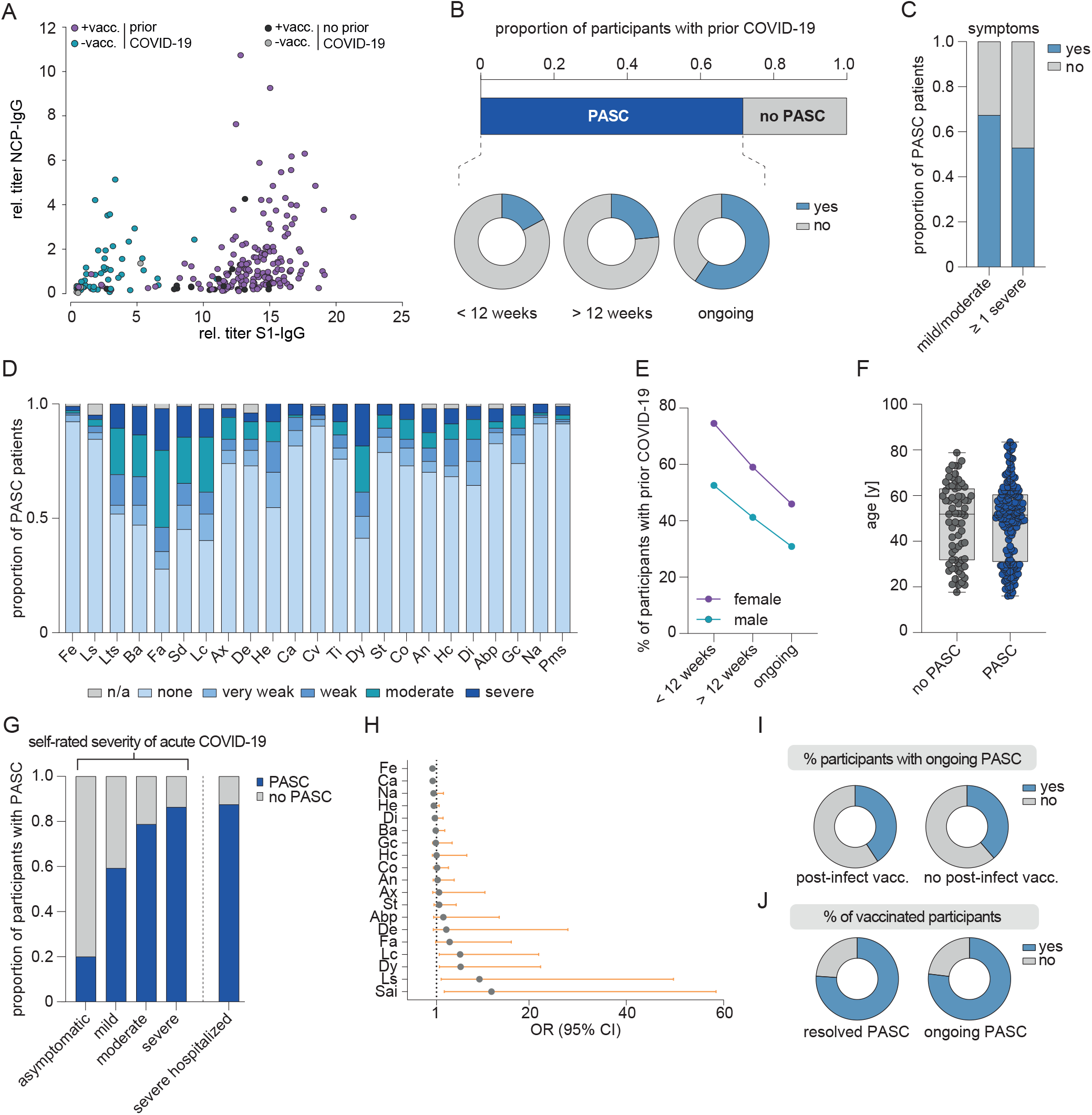
Clinical and epidemiological parameters of the DigiHero cohort and patients with PASC. (A) Plasma titer of antibodies directed against the S1 and NCP proteins of SARS-CoV-2 in individuals with or without SARS-CoV-2 vacciniation (+ vacc./-vacc.) and with or without prior COVID-19 from the DigiHero cohort. (B) Proportion of DigiHero participants with self-reported PASC including duration of PASC symptoms after infection plus proportion of patients with ongoing symptoms at the time of blood sampling. (C) Proportion of PASC patients with mild/moderate or at least one severe symptom. (D) Severity of self-reported symptoms in PASC patients. (E) Distribution of PASC duration between female and male study participants with prior COVID-19. (F) Age distribution of DigiHero participants with or without PASC. (G) Severity of acute COVID-19 in PASC patients. (H) Symptom correlates of acute COVID-19 with PASC shown as odds ration (OR) with 95% confidence interval (CI). (I) Post-vaccination status of patients with ongoing PASC. (J) Vaccination status in participants with resolved or ongoing PASC. Abbreviations: Abp, abdominal pain; An, angina; Ax, anxiety; Ba, body aches; Ca, coryza; Co, cough; Cv, conjunctivitis; De, depression; Di, dizziness; Dy, dyspnea; Fa, fatique; Fe, fever; Gc, gastrointestinal complaints; He, headache; Hc, heart complaints; Lc, lack of concentration; Ls, lymph node swelling; Lts, loss of taste/smell; Na, nausea; Sai, self-reported severity of acute infection; Sd, sleep disturbance; St, sore throat; Ti, tinnitus.

### COVID-19 symptoms and PASC in the DigiHero COVID-19 module

A total of 175 (67.8%) previously infected participants reported symptoms beyond four weeks from positive SARS-CoV-2 testing and were therefore considered to have PASC (Table 1). Distribution of acute COVID-19 and PASC symptoms are shown in Table 2. Of the participants with PASC, 20% showed symptoms only up to 12 weeks and about 60% had ongoing symptoms at the time of blood sampling (Figure 2B). About half of the participants with PASC reported at least one severe symptom beyond four weeks from positive SARS-CoV-2 testing (Figure 2C). Fatigue and dyspnea were amongst the most prevalent symptoms with a considerable fraction of cases showing moderate to severe symptom load (Figure 2D). Women showed a higher percentage of PASC than men and the percentage of patients reporting PASC decreased with time (Figure 2E). PASC was distributed equally across age groups (Figure 2F). Participants with more severe acute infections more likely reported to be affected by PASC (Figure 2G). To evaluate if specific acute COVID-19 symptoms were predictive for PASC, we fitted a generalized linear model. Lymphnode swelling, concentration deficit and dyspnea in acute COVID-19 were significantly associated with PASC (Figure 2H).

**Table 2:**
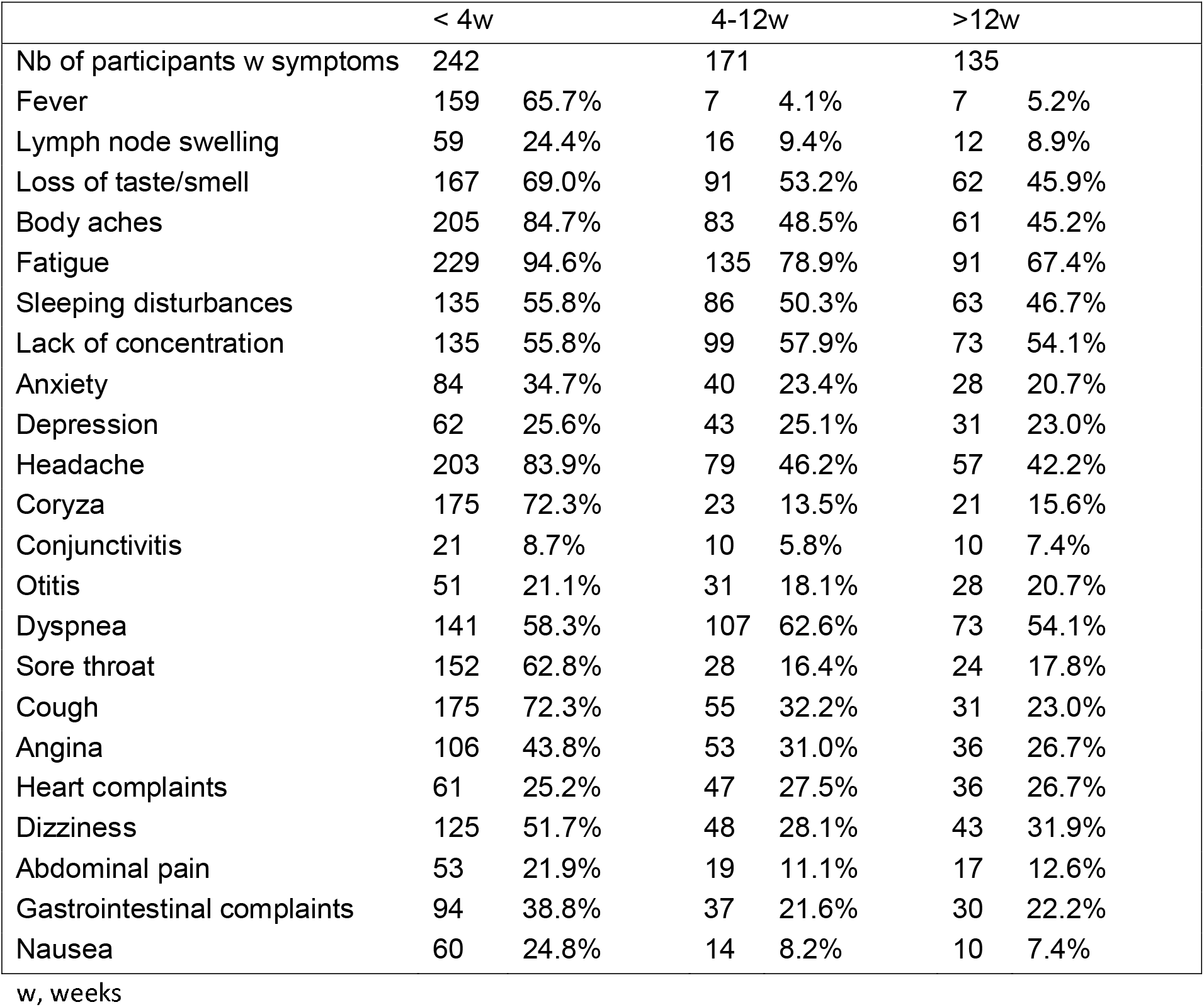
Duration of symptoms.

|  | < 4w |  | 4-12w |  | >12w |  |
| --- | --- | --- | --- | --- | --- | --- |
| Nb of participants w symptoms | 242 |  | 171 |  | 135 |  |
| Fever | 159 | 65.7% | 7 | 4.1% | 7 | 5.2% |
| Lymph node swelling | 59 | 24.4% | 16 | 9.4% | 12 | 8.9% |
| Loss of taste/smell | 167 | 69.0% | 91 | 53.2% | 62 | 45.9% |
| Body aches | 205 | 84.7% | 83 | 48.5% | 61 | 45.2% |
| Fatigue | 229 | 94.6% | 135 | 78.9% | 91 | 67.4% |
| Sleeping disturbances | 135 | 55.8% | 86 | 50.3% | 63 | 46.7% |
| Lack of concentration | 135 | 55.8% | 99 | 57.9% | 73 | 54.1% |
| Anxiety | 84 | 34.7% | 40 | 23.4% | 28 | 20.7% |
| Depression | 62 | 25.6% | 43 | 25.1% | 31 | 23.0% |
| Headache | 203 | 83.9% | 79 | 46.2% | 57 | 42.2% |
| Coryza | 175 | 72.3% | 23 | 13.5% | 21 | 15.6% |
| Conjunctivitis | 21 | 8.7% | 10 | 5.8% | 10 | 7.4% |
| Otitis | 51 | 21.1% | 31 | 18.1% | 28 | 20.7% |
| Dyspnea | 141 | 58.3% | 107 | 62.6% | 73 | 54.1% |
| Sore throat | 152 | 62.8% | 28 | 16.4% | 24 | 17.8% |
| Cough | 175 | 72.3% | 55 | 32.2% | 31 | 23.0% |
| Angina | 106 | 43.8% | 53 | 31.0% | 36 | 26.7% |
| Heart complaints | 61 | 25.2% | 47 | 27.5% | 36 | 26.7% |
| Dizziness | 125 | 51.7% | 48 | 28.1% | 43 | 31.9% |
| Abdominal pain | 53 | 21.9% | 19 | 11.1% | 17 | 12.6% |
| Gastrointestinal complaints | 94 | 38.8% | 37 | 21.6% | 30 | 22.2% |
| Nausea | 60 | 24.8% | 14 | 8.2% | 10 | 7.4% |

w, weeks

### Post-infection vaccination was not associated with resolution of PASC in the DigiHero cohort

Anecdotal reports suggested that SARS-CoV-2 vaccination may lead to resolution of PASC[36] possibly via elimination of a cryptic viral reservoir by the induced immune response. In our cohort, however, we found that the percentage of patients with ongoing PASC was similar in participants with post-infection vaccination and those without (Figure 2I). Moreover, we assessed the percentage of vaccinated participants in two subgroups with PASC. The percentage of post-infection vaccinations was identical in patients with PASC that experienced resolution of their symptoms and in those that showed ongoing PASC (Figure 2J). Together, these data did not lend support to the hypothesis that post-infection vaccination fosters the resolution of PASC.

### Elevated autoantibody levels after COVID-19 were not associated with PASC

We performed autoantibody screens and correlated autoantibody positivity with clinical symptoms. We included rheumatoid factor, antinuclear antibodies and anti-phospholipid antibodies in our assessment[20]. While the control cases without prior COVID-19 infection did not show positivity for any of the tested specificities, the percentage of participants with positive test results for one or more autoantibodies amounted to 20% of prior COVID-19 cases (Figure 3A and B). Interestingly, participants with earlier SARS-CoV-2 infections showed equal rates of autoantibody-positivity compared to participants with later infections arguing against their short-lived nature (Figure 3B). Yet, positivity for these autoantibodies did not correlate with PASC (Figure 3C). Of note, in none of the participants, a new diagnosis of a *bona fide* autoimmune disorder was reported in the surveyed period. Two participants reported worsening of preexisting rheumatoid arthritis or psoriasis, respectively. However, these two cases tested negative for the autoantibodies included in our screening panel. Together, this data does not support an involvement of the tested autoantibody classes in the pathogenesis of PASC.

**Figure 3.**
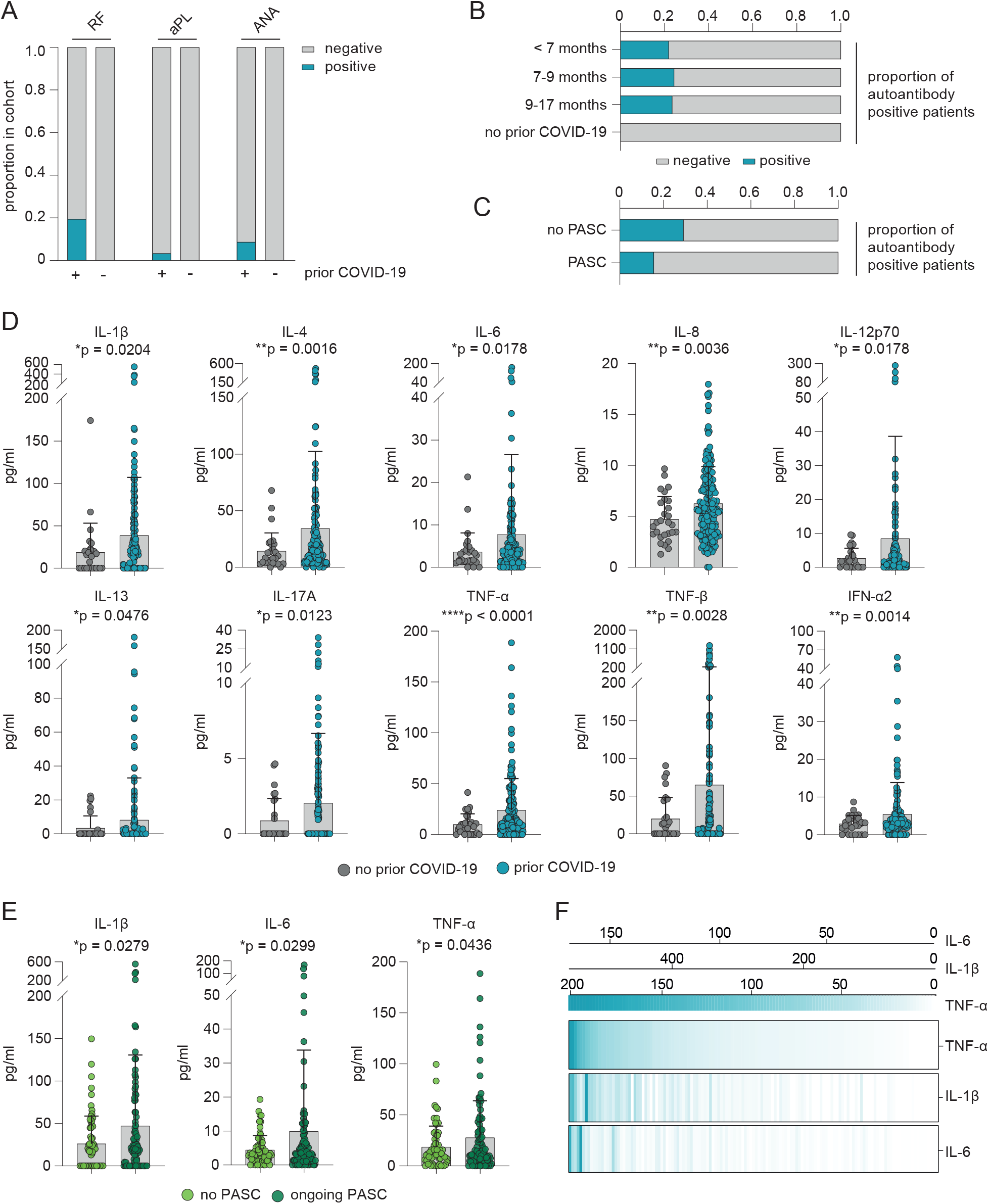
Serological profiling of plasma after resolved SARS-CoV-2 infection and ongoing PASC. (A) Proportion of participants with rheumatic factor (RF), antinuclear (ANA) and phospholipid autoantibodies (aPL) dependent on COVID-19 history. (B) Seroprevalence of autoantibodies over time in COVID-19 patients. (C) Seroprevalence of autoantibodies in the DigiHero cohort and PASC. (D) Mean plasma cytokine levels of participants with or without prior COVID-19. Error bars indicate ± SD. Statistical analysis: two-sided t-test. (E) Mean plasma cytokine levels in patients with or without ongoing PASC at the time of blood sampling. Error bars indicate ± SD. Statistical analysis: two-sided t-test. (F) Correlation of TNF-α, IL-1β and IL-6 plasma levels in patients with ongoing PASC. Concentrations shown as pg/ml.

### Participants with PASC show a pro-inflammatory blood cytokine profile

Next, we profiled plasma samples from participants without prior COVID-19, previously infected participants without PASC and those with ongoing PASC for a broad range of cytokines that are deregulated in acute COVID-19[37]. Despite the long interval between acute infection and blood sampling of eight months, participants with prior COVID-19 showed patterns of systemic cytokine deregulation also found in acute COVID-19 or early recovery[37] including TNF-α, TNF-β, IL-1β, IL-6, IL-8 and IL-12p70 (Figure 3D, Supplementary Figure 1A). Of those, only IL-1ß, IL-6 and TNF-α showed a significant correlation with PASC (Figure 3E, Supplementary Figure 1B). Interestingly, the levels of these three cytokines were positively correlated with each other in individual participants indicating that they do not identify separate subsets of patients with PASC (Figure 3F). These data suggest that persistently elevated levels of IL-1ß, IL-6 and TNF-α may be a hallmark of PASC.

### Single-cell analysis of lung macrophages from COVID-19 patients with acute disease recapitulate cytokine profile found in PASC

While the triad of IL-1ß, IL-6 and TNF-α was characteristic for cases with PASC, the evolution of these cytokines from acute infection to the PASC stage as well as their cellular compartment of origin remained elusive. Since the COVID-19 module of the DigiHero study did not include participants with active COVID-19, we were unable to extrapolate the dynamics of the three PASC-associated cytokines from early infection to post-acute disease phases. To close this gap, we performed plasma cytokine profiling on additional samples from our independent HACO cohort that recruited patients with active infections and in the early post-infection period[37]. Fifteen patients with mild to moderate acute COVID-19 were included that matched the characteristics of the DigiHero cohort (0% versus 1.5% intensive care unit rate). IL-1ß and TNF-α levels were elevated in acute infection with concentrations exceeding those found in patients with bacterial pneumonia (Figure 4A, Supplementary Figure 2). Despite the cross-sectional character of this analysis, this data suggested different dynamics of the three cytokines over time. While IL-6 and TNF-α levels remained essentially stable in the acute and post-acute disease phases, IL-1ß showed a late peak two to three months after infection (Figure 4A).

**Figure 4.**
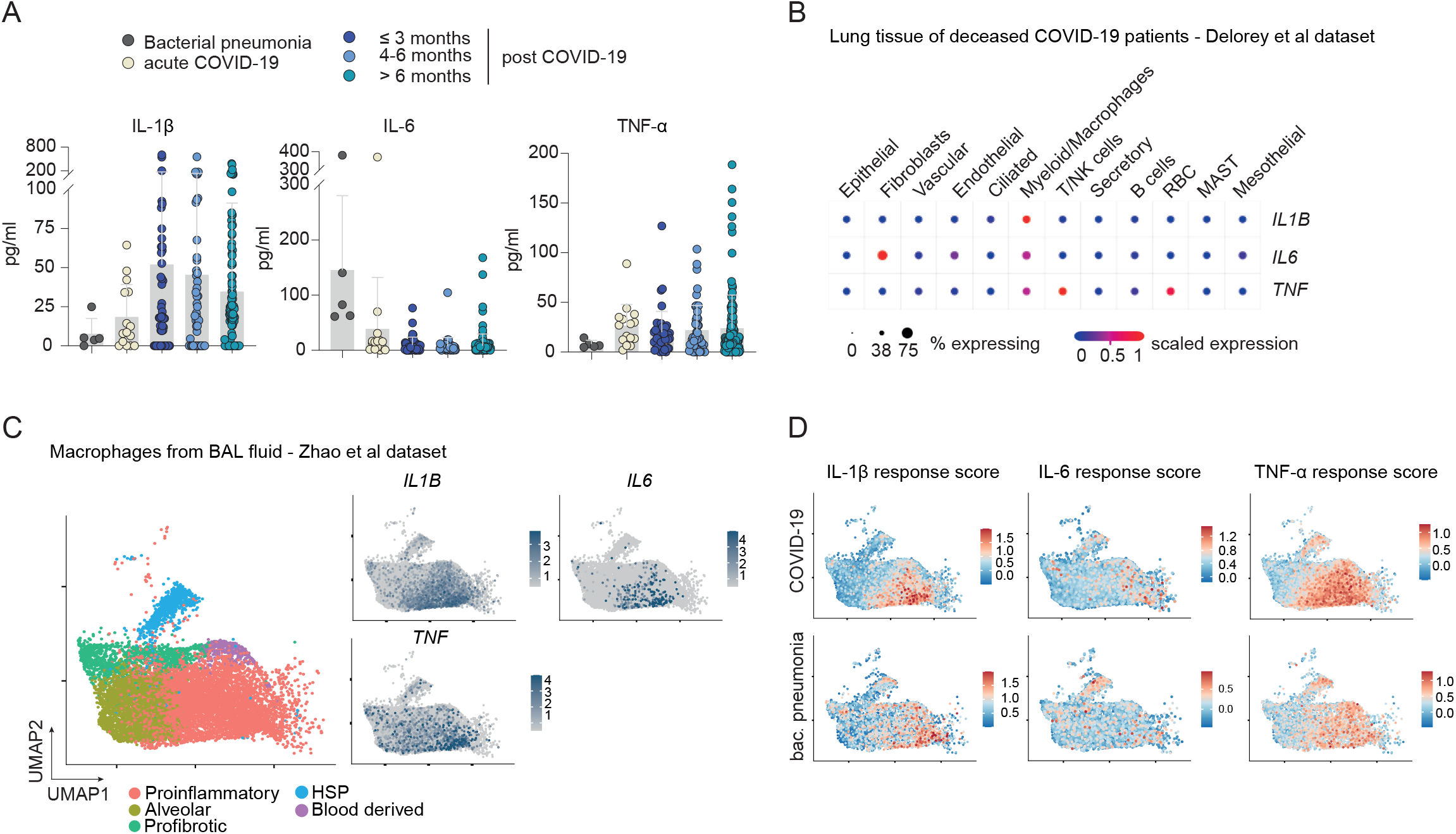
PASC-related cytokine triad in acute COVID-19. (A) Mean plasma levels of TNF-α, IL-1β and IL-6 in acute COVID-19 and their dynamics in post-acute disease phases as compared to patients with bacterial pneumonia. Samples from patients with bacterial pneumonia and acute COVID-19 derived from HACO trial; follow-up blood samples derived from DigiHero and HACO trial. (B) Profiling of *IL1B, IL6* and *TNF* transcripts in lung autopsy tissue from deceased COVID-19 patients. Single-cell transcriptome dataset from [32]. (C) Macrophage subsets from bronchoalveolar (BAL) fluid in active COVID-19. Single-cell dataset from[31]. Uniform Manifold Approximation and Projection (UMAP) plot showing expression of *IL1B, IL6* and *TNF* in macrophage subpopulations. (D) Analysis of gene set associated with response to cytokine triad in macrophage subsets from bronchoalveolar fluid in active COVID-19.

IL-1ß, IL-6 and TNF-α are cytokines which are mainly secreted by monocytes and macrophages upon inflammatory stimuli[38]. The myeloid compartment and especially lung macrophages are strongly deregulated in COVID-19 patients as evidenced by previously published single cell analyzes on lung tissue, bronchoalveolar lavage fluid (BALF) and blood from patients with acute COVID-19[31, 39]. To explore the potential cellular sources of IL-1β, IL-6 and TNF-α in acute COVID-19, we analyzed previously published single-cell transcriptomics datasets from lung tissue, peripheral blood and BALF of patients in acute infection[31-33]. This analysis confirmed that this triad of cytokines was specifically expressed by the myeloid/macrophage compartment in the lungs (Figure 4B), but not or to a lesser extent by the myeloid/monocyte compartment in blood of patients with acute COVID-19 (Supplementary Figure 3). In a next step, we looked in-depth into macrophage subpopulations from bronchoalveolar fluid of patients with acute COVID-19. This analysis showed that the production of IL-1ß, TNF-α and to a lesser extent that of IL-6 was concentrated in the pro-inflammatory subset of lung macrophages (Figure 4C). Gene set analysis covering transcripts of response genes to these cytokines suggested that the pro-inflammatory macrophage not only produced this cytokine triad, but also responded to it (Figure 4D). The inclusion of the late phase TNF response genes *CLU, TNIP3, SIGLEC10, ENPP2* and *NKG7*[34] in the TNF response score further supported a chronic, self-sustaining activation of pro-inflammatory macrophage.

## Discussion

A considerable fraction of patients do not fully recover from COVID-19, but experience lasting sequelae. There are a number of recent population and registration studies addressing the long-term outcomes of patients with COVID-19 both with and without prior hospital admission. These studies suggest that approximately 32-87% of patients infected with SARS-CoV-2 develop PASC[8]. In addition to the inherently restricted follow-up of these studies, their major limitation is the lack of bioimmunological data acquired in subjects with PASC. The wealth of studies dissecting the biology of the acute COVID-19 infection dramatically contrasts with the paucity of biological data currently available for patients with persistent symptoms. As a consequence, current concepts of the natural history of PASC and its pathophysiological drivers remain hypothetical. Yet, this data is urgently needed to develop rational therapeutic strategies.

In the work presented here, we aimed to address these questions by analyzing the first 318 participants from the DigiHero cohort that follows an ambidirectional digital epidemiology approach with a survey focusing on the COVID-19 and vaccination history and a prospective part including an invitation to donate blood for (auto-)antibody and cytokine profiling.

We found that approximately 70% of previously infected participants experienced prolonged symptoms independent of age and were therefore categorized as affected by PASC. Approximately 40% of the initially infected subjects even showed ongoing symptoms at the time of blood sampling which was on average eight months after positive SARS-CoV-2 testing including three individuals with persistent symptoms 16-17 months post-COVID-19 which is substantially longer than the durations reported so far by others[8]. Notably, after other SARS-CoV or MERS infections, long-lasting post-acute sequelae including fatigue, pain and psychiatric morbidities are also common for up to 36 months and reported as chronic post-SARS/MERS syndrome[40-42]. Similar to SARS-CoV and MERS and in line with other studies on SARS-CoV-2[8, 10], the most consistently reported PASC symptoms were fatigue, dyspnea, loss of taste and smell and neurophysiological manifestations including sleep disturbances and lack of concentration. We also found that distinct features of acute COVID-19 were predictive for development of PASC, including lymphnode swelling, concentration deficit and dyspnea as well as the severity of acute disease. While the predictive value of severity of acute COVID-19 was also acknowledged by others[10], it is noteworthy that severity in our cohort was self-reported and not associated with hospitalization or ICU admission as in other studies. This suggests that PASC may also arise in a substantial proportion of infected individuals who do not require medical intervention during active disease.

One hypothetical mechanistic explanation for PASC could be inefficient clearing of SARS-CoV-2 resulting in cryptic viral reservoirs especially outside the lung. In this notion, vaccination-induced boosting of the SARS-CoV-2-directed immune response towards increased neutralizing breadth and potency could contribute to eradication of latent virus or its immunogenic remnants and thereby to the resolution of PASC. Indeed, while SARS-CoV-2 RNA shedding peaks around 10-14 days post infection, persistence in serum and stool samples up to 126 days post infection has been reported - similarly to SARS-CoV and MERS[43, 44]. In addition, viral RNA and protein were detected in the lower gastrointestinal tract in about 30% of tested individuals with negative nasopharyngeal-swab PCR up to 6 months post-infection[16]. Our digital cohort allowed us to assess the relationship between post-infection vaccination and resolution of PASC. These data show that the post-infection vaccination rate is equally high in participants in which PASC eventually resolves as in those with ongoing PASC. Moreover, the percentage of participants with ongoing PASC was comparable in vaccinated and non-vaccinated subgroups. This data does not *prima facie* support the hypothesis of a cryptic SARS-CoV-2 reservoir underlying PASC that may be eradicated by a vaccination-boosted immune response.

A striking feature of COVID-19 are systemic manifestations of autoimmunity mirrored by elevated levels of different autoantibody classes[20-26], a feature that is shared by many viral infections including SARS-CoV and Mers[45, 46]. While autoantibody positivity correlates with COVID-19 severity, the potential pathophysiological relevance for PASC is unclear. In our cohort, there was no detectable correlation of autoantibodies with PASC as well as no evidence for newly emerging autoimmune conditions. The clinical relevance of such SARS-CoV-2-induced autoantibodies therefore still remains debatable.

In contrast, our cytokine profiling revealed a significant association of a well-known triad of cytokines - IL-1ß, IL-6 and TNF-α - with PASC. Due to their functional role in pain perception, anxiety, depression and inflammatory symptoms[47-56] they are well compatible with the spectrum of symptoms in PASC. Analysis of macrophages from the bronchoalveolar lavage fluid in patients with severe COVID-19 showed that a specific pro-inflammatory macrophage subset produced high levels of these cytokines in acute disease. Mining of this single-cell sequencing dataset for cytokine response pathways revealed that such macrophages may not only be primed in the lung to produce the cytokine triad, but may also respond to it. The nature of response genes involved suggested chronic cytokine exposure. Based on this data, one may speculate that acute pro-inflammatory reprogramming of long-lived lung macrophages or their precursors may result in a vicious circle of IL-1ß, IL-6 and TNF-α production that self-maintains this cellular compartment. This hypothesis may be supported by recent evidence showing epigenetic reprogramming of the monocyte/macrophage compartment in COVID-19 that results in pathological inflammasome engagement in these cells[57]. Yet, to definitively prove the causal relationship between the cytokine triad and self-sustained, chronic activation of reprogrammed macrophage in patients with PASC, matched tissue and blood analyzes at the PASC stage would be instrumental.

Taken together, the combination of digital epidemiology and selective biobanking put us in a position to rapidly explore biomarkers of PASC in a well characterized cohort of patients with a considerable follow-up of eight months. It is noteworthy, that - owing to the digital recruitment approach - the recruitment time was only two weeks from invitation. Since participants answered the questionnaires online, identification of eligible participants for the COVID-19 module was possible in real-time. Digitalization may therefore considerably accelerate epidemiological health research which is of use not only for pandemic research questions.

## Data Availability

All data produced in the present study are available upon reasonable request to the authors

## Acknowledgement

The DigiHero study is conducted by a consortium of the Medical Faculty of the Martin-Luther-University Halle-Wittenberg including the following PIs: Mascha Binder, Thomas Frese, Michael Gekle, Matthias Girndt, Jessica Höll, Patrick Michl, Rafael Mikolajczyk, Matthias Richter and Daniel Sedding. We thank Aline Patzschke, Christoph Wosiek and Bianca Gebhardt for excellent technical assistance. We sincerely thank healthy donors, patients and their household members for participating in this study. Flow cytometry was performed at the UKH FACS sorting core facility. This project was partially funded by the CRC 841 of the German Research Foundation (to MB) as well as by the Medical Faculty of the Martin-Luther-University Halle (Saale).

## Author contributions

M. Binder, R. Mikolajczyk, M. Gekle, C. Schultheiß, L. Paschold, S. Henkes, C. Gottschick, B. Klee, M. Girndt, T. Frese, D. Sedding and J. Höll designed the COVID-19 module of the DigiHero cohort study. D. Sedding and J. Dutzmann provided the HACO patient cohort. C. Schultheiß and L. Paschold conducted experiments. M. Binder, C. Schultheiß, L. Paschold, E. Willscher and L. Bosurgi analyzed and interpreted the data. M. Binder, C. Schultheiß, E. Willscher and L. Paschold drafted the manuscript. All authors critically revised and approved the manuscript.

## Declaration of Interests

The authors declare no competing interests.

## Figure Legends

**Supplementary Figure 1. Cytokine levels in plasma of individuals with or without prior COVID-19 and with or without ongoing PASC**.

(A) Mean plasma cytokine levels of participants with or without prior COVID-19. Error bars indicate ± SD. Statistical analysis: two-sided t-test.

(B) Mean plasma cytokine levels in patients with or without ongoing PASC at the time of blood sampling. Error bars indicate ± SD. Statistical analysis: two-sided t-test.

**Supplementary Figure 2. PASC-related cytokine triad in acute COVID-19**.

Mean plasma levels of indicated cytokines in acute COVID-19 and their dynamics in post-acute disease phases as compared to patients with bacterial pneumonia. Samples from patients with bacterial pneumonia and acute COVID-19 derived from HACO trial; follow-up blood samples derived from DigiHero and HACO trial. Error bars indicate ± SD.

**Supplementary Figure 3. Profiling of *IL1B, IL6* and *TNF* in PBMCs of hospitalized COVID-19 patients**.

Expression of the *IL1B, IL6* and *TNF* transcripts in distinct cellular PBMC subsets shown as violin plots. Dataset from [33].

## Notes

### Competing Interest Statement

The authors have declared no competing interest.

### Funding Statement

This study was partially funded by the CRC 841 of the German Research Foundation (to MB) as well as by the Medical Faculty of the Martin-Luther-University Halle (Saale).

### Author Declarations

Ethics committe of Martin-Luther University gave ethical approval of this work (approval numbers 2020-076)

