## Supplemental Figure 1 for "From online data collection to identification of disease mechanisms: The IL-1ß, IL-6 and TNF-α cytokine triad is associated with post-acute sequelae of COVID-19 in a digital research cohort"

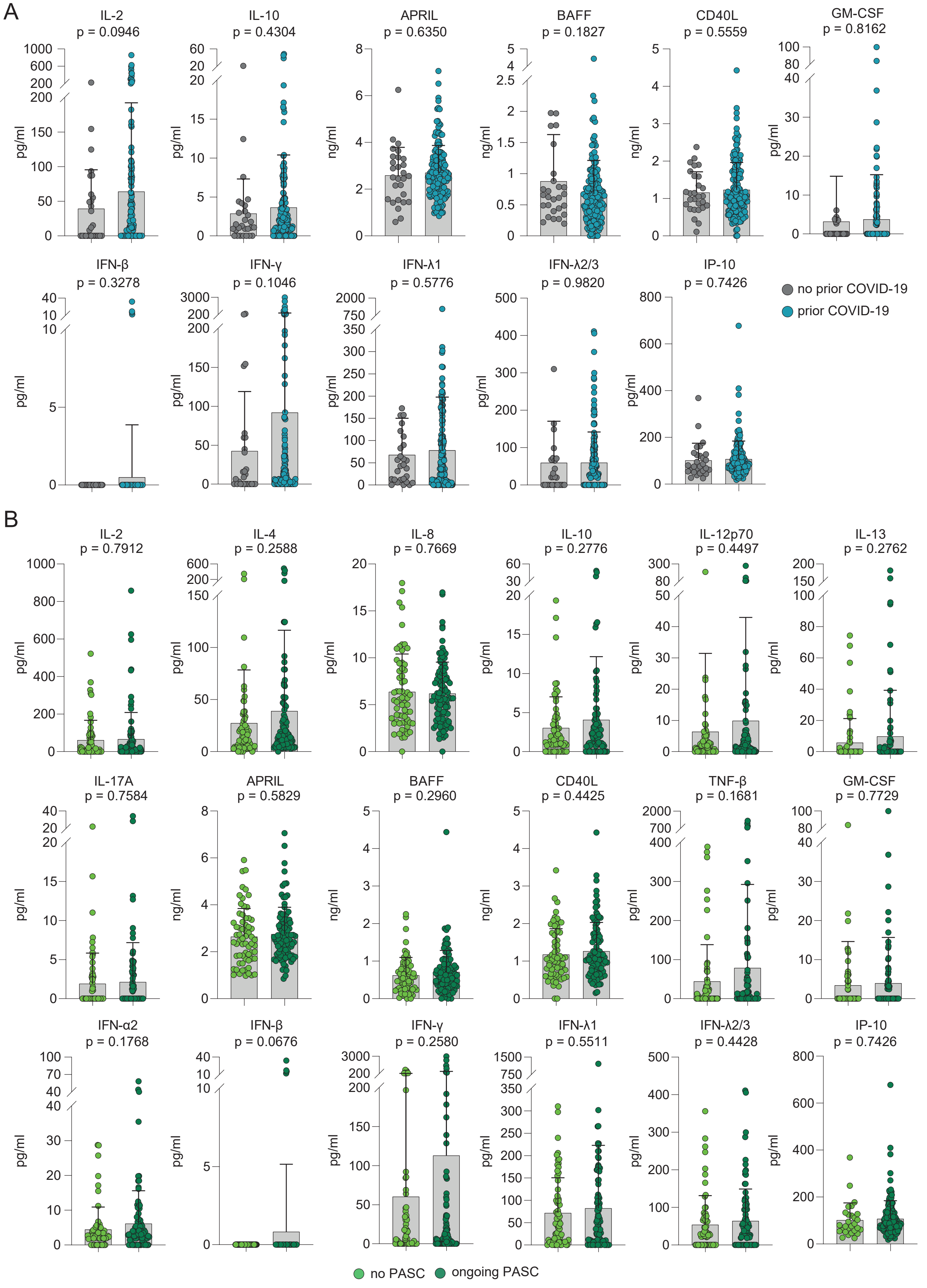


**Supplementary Figure 1. Cytokine levels in plasma of individuals with or without prior COVID-19 and with or without ongoing PASC.**
(A) Mean plasma cytokine levels of participants with or without prior COVID-19. Error bars indicate ± SD. Statistical analysis: two-sided t-test.
(B) Mean plasma cytokine levels in patients with or without ongoing PASC at the time of blood sampling. Error bars indicate ± SD. Statistical analysis: two-sided t-test.
