## Supplemental Figure 2 for "From online data collection to identification of disease mechanisms: The IL-1ß, IL-6 and TNF-α cytokine triad is associated with post-acute sequelae of COVID-19 in a digital research cohort"

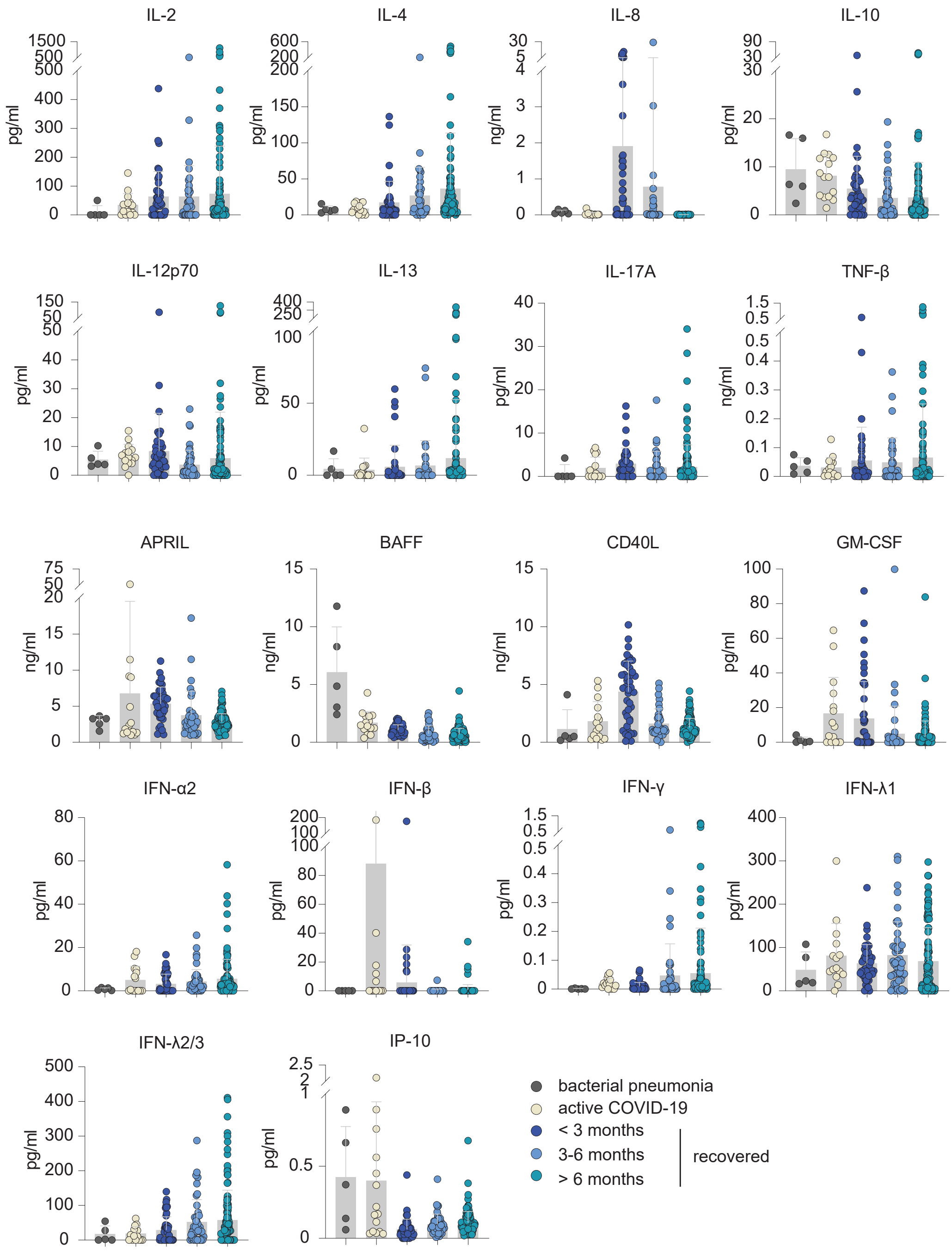


**Supplementary Figure 2. PASC-related cytokine triad in acute COVID-19.** Mean plasma levels of indicated cytokiens in acute COVID-19 and their dynamics in post-acute disease phases as compared to patients with bacterial pneumonia. Samples from patients with bacterial pneumonia and acute COVID-19 derived from HACO trial; follow-up blood samples derived from Digihero and HACO trial. Error bars indicate ± SD.
