## Supplemental Figure 3 for "From online data collection to identification of disease mechanisms: The IL-1ß, IL-6 and TNF-α cytokine triad is associated with post-acute sequelae of COVID-19 in a digital research cohort"

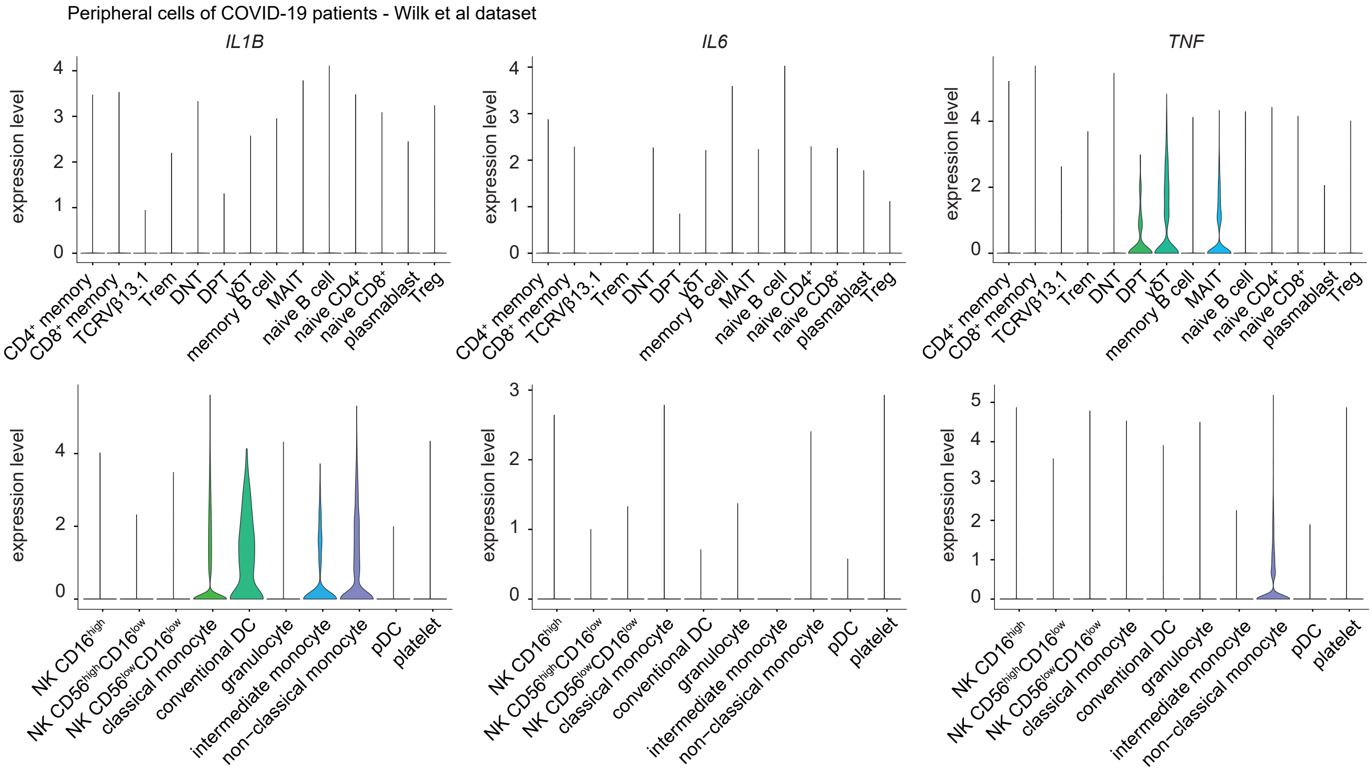


**Supplementary Figure 3. Profiling of IL1B, IL6 and TNF in PBMCs of hospitalized COVID-19 patients.**

Expression of the IL1B, IL6 and TNF transcripts in distinct cellular PBMC subsets shown as violin plots. Dataset from (Wilk et al., 2020).
